# SEROPREVALENCE OF HEPATITIS B VIRUS INFECTION AMONG HIV INFECTED INDIVIDUALS IN UYO, AKWA IBOM STATE, NIGERIA

**DOI:** 10.1101/2021.03.06.21253060

**Authors:** Hope C. Innocent-Adiele, Baah B. T. Michael, Iheanyi O. Okonko, Ogbonnaya Ogbu

## Abstract

**Aim:** Hepatitis B and Acquired Immunodeficiency Syndrome (AIDS) are highly endemic in Nigeria and are important causes of death and disability. Co-infection between hepatitis B virus (HBV) and Human Immunodeficiency Virus (HIV) commonly occur as both viruses share a common mode of transmission. This leads to fulminant hepatitis and liver cirrhosis depending on the stages of infection which are acute and chronic stages respectively. This study was carried to determine the prevalence of hepatitis B virus (HBV) among HIV-infected individuals in Uyo, Akwa Ibom State, Nigeria.

**Methods:** In this study 176 HIV-infected individuals were recruited comprising 67 males and 109 females. These subjects were screened for the presence of hepatitis B surface antigen (HBsAg) using enzyme linked immunosorbent assay.

**Results:** From those tested, 11 were positive for HBsAg giving an overall prevalence rate of 6.3%. Co-infection rate of males (8.5%) did not differ significantly (p>0.05) from that of females (6.4%). Co-infection was highest in age group 6-30 years (28.2%). In relation to marital status, singles had the higher co-infection rates (10.5%) than married subjects (4.7%). Among the different occupational groups, students had the highest co-infection rate (22.2%) and was closely followed by business (16.7%). Higher HIV/HBV co-infection was observed among those with CD4 cell count <200 cells/μl (15.4%) and those with plasma viral loads (PVL) ≥5001 copies/mL (13.6%).

**Conclusion:** This study confirms the high HIV/HBV co-infection rate (6.3%) and thus, there is a need to screen all HIV-positive individuals for HBV infection. A high seroprevalence of HBV among this cohort of HIV-infected individuals contributes to the calls for pre-ART screening for HBV and the necessary paradigm shift in the ART nucleoside backbone to include agent(s) more dually effective against HIV and HBV.

## 1.1 INTRODUCTION

Human immunodeficiency virus (HIV) and hepatitis B virus (HBV) are causes of significant morbidity and mortality across the World (WHO, 2014; 2015). HIV and HBV are blood-borne viruses transmitted mainly through sexual contact and use of unsterilized needles. Their similar means of transmission increases the risk of contracting both infections concurrently (WHO, 2020). Viral hepatitis is an acute or chronic inflammation of the liver caused by viral infection (CDC, 2019). HBV is globally the leading cause of death due to liver disease in people living with HIV/AIDS (Opaleye et al., 2017). In 2016, the World Health Organization (WHO) estimated that 36.9 million people are living with HIV (WHO, 2016). It was also reported that 248 million people have chronic HBV infection (Schweitzer et al., 2015; Katamba et al., 2020). It is estimated that chronic HBV infection affects an estimated 5–20% of people living with HIV (WHO, 2020). HIV-positive persons who become infected with HBV or hepatitis C virus (HCV) are at increased risk for developing chronic hepatitis (WHO, 2020). In addition, persons who are co-infected with HIV and hepatitis can have serious medical complications, including an increased risk for liver-related morbidity and mortality, chronic hepatitis, cirrhosis and hepatocellular carcinoma, all of which are of serious public health concern (Balogun et al., 2012; WHO, 2020).

HIV and HBV are both endemic in Nigeria. There is a heavy burden of HIV – HBV and HIV – HCV co-infections in many regions of the developing world (Cooper et al., 2009), including Nigeria (Ola et al., 2002; Forbi et al., 2007; Balogun et al., 2012). The burden of these co-infections is greatest in the African and South-East Asian regions (WHO, 2020). HIV is associated with a higher prevalence of both HBV and HCV in Sub-Saharan Africa. In this region, many people living with HIV are co-infected with HBV or HCV (Moges et al., 2006; Barth et al., 2010; Shimelis et al., 2017; Katamba et al., 2020). In Nigeria, hepatitis co-infection with HIV is associated with increased morbidity and mortality (Balogun et al., 2012). Nigeria has the second largest HIV prevalence in the world and one of the highest rates of new infection in sub-Saharan Africa (NACA, 2017; AVERT, 2020). Unprotected heterosexual sex accounts for 80% of new HIV infections in Nigeria, with the majority of remaining HIV infections occurring in key affected populations such as sex workers (NACA, 2015). Akwa Ibom State tops the prevalence rate chart with about 5.6% of its residents living with the virus (NAIIS, 2019) and it is one of the six states in Nigeria that accounted for 41% of people living with HIV in Nigeria (NACA, 2017).

The notable relationship between HIV and HBV is that they can be transmitted in the same way. Several studies strongly suggest that the influence of HIV on HBV is characterized by a chronic infection, an increased viral replication rate, higher viral levels, accelerated liver damage and an increased risk of liver cancer (Salmon-Ceron et al., 2005; Cooper et al., 2009; Thio, 2009; Ymele et al., 2012; Stabinski et al., 2015). On the other hand, the HBV infection aggravates the progression of HIV towards AIDS and an increased *in vitro* replication of HIV (Burnett et al., 2005; Ymele et al., 2012). Mortality among HIV/HBV co-infected persons is substantially higher than among HIV mono-infected persons (Fernández-Montero and Soriano, 2012; Mocroft et al., 2012; Teira and VACH Study Group, 2013; Kruse et al., 2014; Stabinski et al., 2015). A study in the USA, revealed that HIV/HBV co-infection increased mortality by 8-fold or 19-fold when compared with single infection with HIV or HBV alone respectively (Thio et al., 2002; Stabinski et al., 2015).

The exact impact of HBV on HIV disease progression is less clear (Hoffmann et al., 2009; Chun et al., 2012; Stabinski et al., 2015), although it is postulated that HBV infection may potentially lead to a blunted immune response in patients receiving antiretroviral therapy (ART) and increase patient susceptibility to ART-related liver toxicity (Wandeler et al., 2013; Stabinski et al., 2015). Unfortunately, access to anti-viral medication is limited in resource-poor countries and only 41% of people living with HIV have access to ART in sub-Saharan Africa (WHO, 2011; Ymele et al., 2012). The objective of this study was to determine the prevalence of hepatitis B virus (HBV) among HIV-infected individuals in Uyo, Akwa Ibom State, Nigeria.

### 2.1 Study Area

The study was conducted in Uyo, Akwa Ibom State, Nigeria. Akwa Ibom is one of the 36 states in Nigeria, with a population of over five million people. The state’s capital, Uyo, has over 500,000 inhabitants and is located in the coastal southern part of the country, lying between latitudes 4°32′N and 5°33′N, and longitudes 7°25′E and 8°25′E. The state is located in the South-South geopolitical zone, and is bordered on the east by Cross River State, on the west by Rivers State and Abia State, and on the south by the Atlantic Ocean and the southernmost tip of Cross River State.

### 2.2 Study design

This is a cross-sectional study carried out in University of Uyo Teaching Hospital (UUTH), Uyo, Akwa Ibom State, Nigeria. HIV antigen detection and serological analyses for hepatitis B surface antigen (HBsAg) were conducted at the Virus Research Unit of the Department of Microbiology, University of Port Harcourt. Port Harcourt, Nigeria.

### 2.3 Ethical Considerations

Approval to undertake the study was obtained from the University Research Ethics Committee of the University of Uyo Teaching Hospital (UUTH), Uyo, Akwa Ibom State, Nigeria. A formal informed consent form was received from each participant before commencement of the study. A healthcare officer of the hospital who is knowledgeable in the care and support of people living with HIV/AIDS assisted with the administration of the informed consent forms, obtaining patients’ demographic data and other pertinent information within the administered questionnaires.

### 2.4 Study population

The study population included male and female individuals living with HIV that attend clinic at the University of Uyo Teaching Hospital (UUTH). One hundred and seventy-six (176) HIV-infected individuals were selected and participated in the study.

### 2.5 Inclusion and exclusion Criteria

Individuals that were included in the study were males and females confirmed and documented as being positive for HIV infection. These selected individuals had or did not have a drug history of anti-retroviral therapy (ART). While individuals who decline involvement in the study were not included in the study.

### 2.6 Data collection

A random sampling irrespective of age, gender and ethnicity was done to ensure that sampling was representative of Uyo, Akwa Ibom State, Nigeria. Necessary demographic (age, sex, marital status, education and occupation), clinical, and epidemiological data for every participant was obtained using a well-structured questionnaire.

### 2.7 Sample Collection and Preparation

After obtaining written informed consents from the participants, blood samples (about 5ml) were aseptically collected during routine investigations so that the participants were not bled twice. The samples were collected into sterile EDTA bottles and plasma samples were obtained after centrifugation. Samples were appropriately labelled and stored in two aliquots at −20°C and −80°C until analysis.

### 2.8 Serological analysis

#### 2.8.1. Rapid Assay for Hepatitis B Surface Antigen (HBsAg)

The assay was carried out using DiaSpot HBsAg rapid test strip (DiaSpot Diagnostics, USA) and interpreted according to the manufacturer’s specifications. The test strip is a rapid, one step test for the qualitative detection of HBsAg in serum or plasma. The test strip uses the immuno-chromatographic method to detect the presence or absence of HBsAg in serum or plasma. All test strips, serum or plasma specimens, and controls were allowed to equilibrate to room temperature (15-30°C) prior to testing. The assay was performed within 1 hour in order to obtain best results according to the manufacturer’s specifications. The result was read at 15 minutes and no result was interpreted after 30 minutes because a low HBsAg concentration might result in a weak line appearing in the test region (T) after an extended period of time. The interpretation of test results was performed according to the manufacturer’s specifications.

#### 2.8.2. ELISA for Detection of Hepatitis B Surface Antigen (HBsAg)

Serum samples were analyzed for hepatitis B surface antigen (HBsAg) using the ELISA kit (DIA.PRO Diagnostic Bioprobes, Italy). The tests were performed according to the manufacturer’s instructions. The test results were calculated by means of a cut-off value determined on the mean OD450nm value of the negative control (NC) with the following formula: NC + 0.050 = Cut-Off (Co). Test results are interpreted as ratio of the sample OD450nm (S) and the Cut-Off value (Co), mathematically S/Co, according to the following: < 0.9 = negative, 0.9 – 1.1 = equivocal and > 1.1 = positive. A negative result indicated that the patient is not infected by HBV. A positive result was indicative of HBV infection and therefore the patients should be treated accordingly.

### 2.9 CD4 T cell count enumeration

EDTA-treated blood samples were used for CD4 T cell count using Partec CyFlow^®^ Counter (Partec GmbH, Germany), following the instruction of manufacturer. The specimens were analyzed on a flow cytometer for detection of cell surface markers for CD4 cells. Results were classified based on the CDC (1997) guidelines.

### 2.10 HIV-1 Viral Load Testing (Abbott Real-Time Assay)

To assess the viral load (VL) of HIV-1 positive individuals used in this study, Abbott RealTime HIV-1 (m2000sp) assay was used to determine the viral load according to the manufacturers’s instruction. The results were presented in copies/mL of plasma.

### 2.11 Statistical analysis of data

Generated data from the study were presented with descriptive statistics (number with percentage; mean; standard deviation or median with range etc.). Data analysis was carried out and test of significance was done using Microsoft Excel 2016 version. Differences of P<0.05 was taken to be statistically significant.

## 3. RESULTS

### 3.1 General characteristics of HIV-infected individuals

As shown in Table 1, 176 (42.2%) from Akwa Ibom State, Nigeria. The age range of the 176 HIV-1 positive individuals who participated in the study was 6-72 years with an average age of 40.0 years. About 56.8% of them were in the 31-40 years age range. The majority (61.9%) of the HIV-1 infected individuals were females and 38.1% were males (Table 1).

**Table 1:**
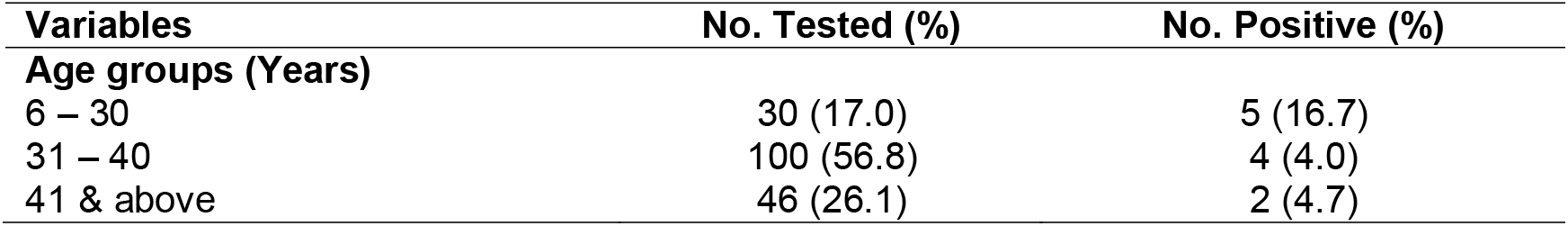
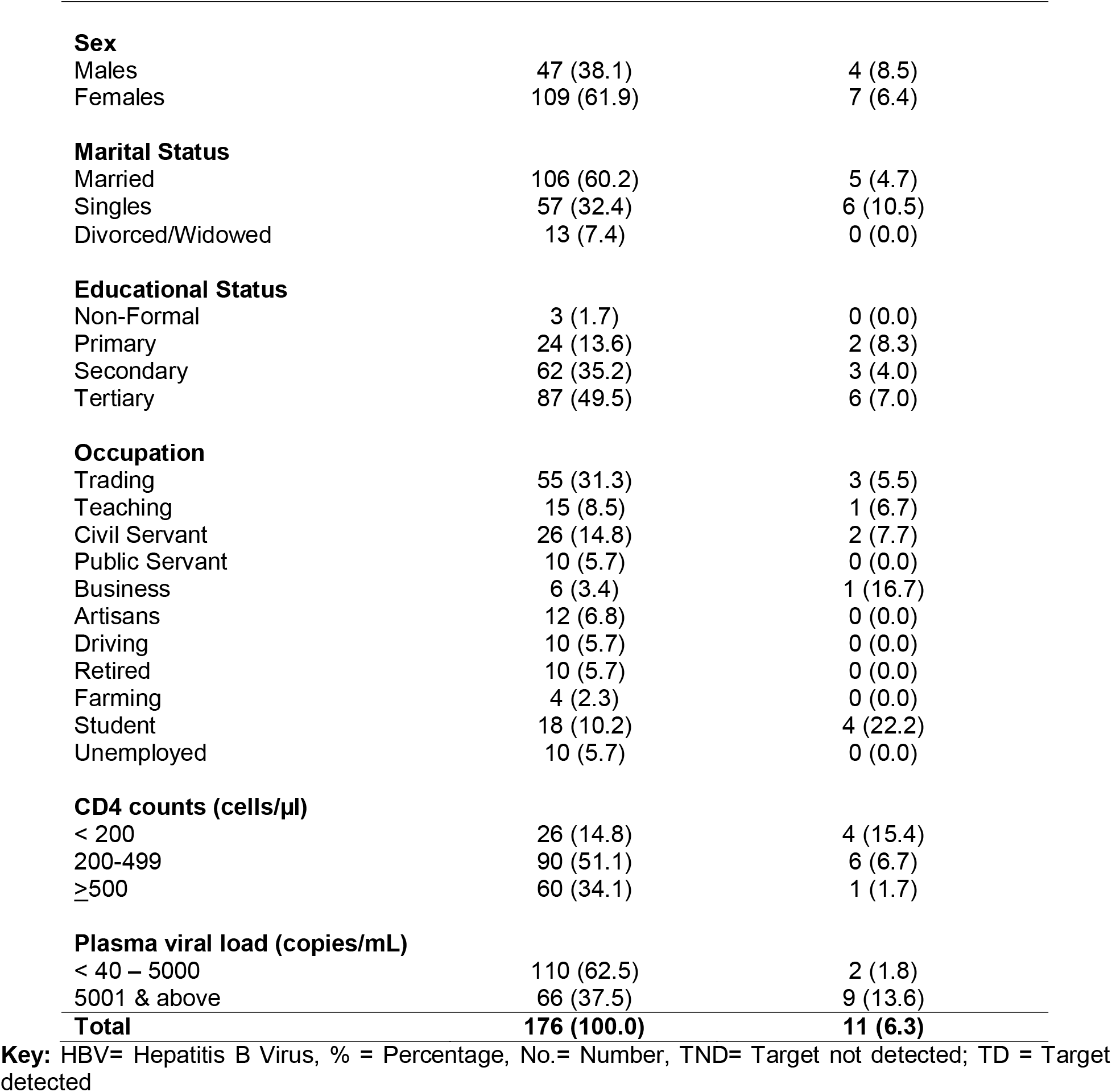
Socio-demographic characteristics of HIV-infected individuals with HIV/HBV co-infection in Uyo, Nigeria.

### 3.2 Overall prevalence of HBsAg in HIV-infected individuals in Uyo, Nigeria

From Table 1, it was shown that 11 out of the 176 samples analyzed for hepatitis B surface antigen (HBsAg) tested positive, giving rise to a prevalent rate of 6.3%.

### 3.3 Age-specific prevalence of HBsAg among HIV-infected individuals in Uyo, Nigeria

The age-specific prevalence of HBsAg among HIV-infected individuals was higher in the age group of 6-30 years (28.6%) than in other age groups (Table 1).

### 3.4 Sex-specific prevalence of HBsAg among HIV-infected individuals in Uyo, Nigeria

The sex-specific prevalence of HBsAg among HIV-infected individuals was higher in males (8.5%) than in females (6.4%) (Table 1).

### 3.5 Marital status-specific prevalence of HBsAg among HIV-infected individuals in Uyo, Nigeria

The marital status-specific prevalence of HBsAg among HIV-infected individuals was higher in singles (10.5%) compared to the married (4.7%) (Table 1).

### 3.6 Education-specific prevalence of HBsAg among HIV-infected individuals in Uyo, Nigeria

The education-specific prevalence of HIV-HBV co-infections in this study revealed a higher prevalence among primary educational status (8.3%) compared to other educational status (tertiary 6.9% and secondary 4.9%) (Table 1).

### 3.7 Occupation-specific prevalence of HBsAg among HIV-infected individuals in Uyo, Nigeria

The occupation-specific prevalence of HBsAg among HIV-infected individuals was higher in students than any other variables in relation to occupation with a prevalence rate of 22.2% (Table 1).

### 3.8 CD4 count-specific prevalence of HBsAg among HIV-infected individuals in Uyo, Nigeria

Higher HIV/HBV co-infection was observed among HIV-infected individuals with CD4 T cell count <200 cells/μl (15.4%) compared to 200 −499 cells/μl (6.7%) and ≥500 cells/μl (1.7%) (Table 1).

### 3.9 Plasma viral load-specific prevalence of HBsAg among HIV-infected individuals in Uyo, Nigeria

Higher HIV/HBV co-infection was observed among HIV-infected individuals with plasma viral loads (PVL) 5001 copies/mL and above (13.6%) than those with <40-5000 copies/mL (1.8%) (Table 1).

## 4. DISCUSSION

Globally, it is estimated 5%–10% of people living with HIV are co-infected with hepatitis B virus (HBV) while HIV/HBV frequency in sub-Saharan Africa varied from 0.0% to 28.4% (Stabinski et al., 2015). HIV and HBV share common risk factors, and many cases of HIV occur in people with HBV, resulting in an increased risk for HIV/HBV co-infection (Hoffmann and Thio, 2007). Nigeria is also known to be highly endemic for Hepatitis B viral (HBV) infection (Anigilaje and Olutola, 2013). There is a relative paucity of data on HIV/HBV co-infection. Our study examined the prevalence of HBsAg among people living with HIV in Uyo, Nigeria. We observed HBsAg positivity of 11 (6.3%) among HIV infected individuals. The co-infection rate of 6.3% reported in this study is a clear indication due to the fact that HBV is a major threat to HIV/AIDs patients in Nigeria. It is comparable to some previous studies that have revealed 6.0% as prevalence of HIV/HBV co-infection in South Africa and in Greece (Lodenyo et al., 2000; Ramezani et al., 2009). It is higher than 2.2% documented in south-western part of Nigeria (Ajayi *et al*., 2013), and 5.8% reported in south eastern region of Nigeria (Nwolisa *et al*., 2013), but lower than the 15.0% reported in north central region of Nigeria (Adoga *et al*., 2009). The factors driving these regional differences are unclear.

The 6.3% reported in this study is lower than the values reported in other countries of the world. A study in the Netherlands identified that 3.6% (Bloquel et al., 2010) of HIV-infected patients were HBsAg positive. It is lower than 4.7% (Rahimi *et al*., 2009) obtained from a study in Spain. This study was also found to be higher than 3.7% obtained in a study in Brazil (Tounkara et al., 2009), 3.6% in Netherlands (Bloquel et al., 2010), 0.24% in Enugu (Ikeako et al., 2014), 0.7% in Anambra State (Ezegbudo et al., 2004), 0.78% in Cameroon (Ymele et al., 2012), 1.4% in United States (Kruse et al., 2014), 4.9% in Canada (Moradi *et al*., 2011), 5.4% in France (Nordenstedt *et al*., 2010), 4.5% in South America (Almeida *et al*., 2006), 4.7% in Spain (Rahimi-Movaghar et al., 2009), and 3.2% in Japan (Rai *et al*., 2007). The value reported in this study is not comparable with reports by Forbi et al. (2007) in North Central Nigeria, South Africa (Parboosing et al., 2008), Senegal (Diop-Ndiaye et al., 2008) and France (Larsen et al., 2008). This study was also found to be lower than the 12.1% prevalence obtained in a study in Burkina Faso (Lodenyo *et al*., 2000), and 8.7% in Thailand (Sungkanuparph et al., 2004), 7.3% in India (Osborn *et al*., 2007), 7.8% in France (Landes et al., 2008), 9.4% in Germany (Aparicio et al., 2012), 15.4% in Italy (Bloquel et al., 2014), 45.0% in France (Nikolopoulos et al., 2009), 59.0% in Finland (Piroth et al., 2008), 27.0% in China (Anggorowati *et al*., 2012), 14.5% in Iran (Fujisaki et al., 2011), 11.3% in Iran (Tsuchiya et al., 2012), 7.8% in Iran (Babamahmoodi et al., 2012), and 28.6% in another study in Iran (Liang et al., 2010).

The observed HIV/HBV co-infection rate (6.3%) in this study is not comparable with previous studies which reported high figures in different parts of Nigeria and overseas; 28.4% in Lagos (Balogun et al., 2012), 20.6% in Keffi (Forbi et al., 2007), 28.7% in Jos (Irisena et al., 2002), 30.4% in Ilorin (Olatunji and Iseniyi, 2008), 70.5% in Kano (Nwokedi et al., 2006), 33.8% in India (Sud et al., 2001), 9.5%, 9.9% and 12.2%, respectively, in Zambia (Katamba et al., 2020; Kapembwa et al., 2011; Vinikoor *et al*. (2015). It is also lower than results obtained by Mbanya et al. (2003) in 2001 and Laurent et al. (2010) at Yaounde, Cameroon. Indeed, Laurent et al. (2010) found among HIV-positive patients, 8.3% co-infections with HBV. Some previous reports of HIV/HBV co-infection from Nigeria, 26.5% and 10.4% among HIV-infected and non-HIV infected in Gombe (Mustapha and Jibrin, 2004), 15.0% in Maiduguri (Baba et al., 1998), 9.2% in Lagos (Lesi et al., 2007), and 9.7% Niger Delta (Ejele et al., 2004) are however higher than observed figures in this study.

The prevalence HIV/HBV co-infections of males (8.5%) in this study is higher than that of females (6.4%), but this was not statistically significant. On the contrary, Mustapha and Jibrin (2004) reported higher rate in females (28.2%) than males (24.7%), Ymele et al. (2012) reported higher rate in females (1.30%) than males (0.74%), and that of Anigilaje and Olutola (2013) also reported that more female subjects (10.0%) were dually infected with HIV and HBV than male subjects (5.9%). However, the findings of this present study are comparable to that of another study we conducted in Rivers State, Nigeria (Okonko et al., 2020) and a study conducted in North Western Nigeria which found that males with higher prevalence (16.9%) compared to females (9.2%) (Muhammad *et al*., 2013). Balogun et al. (2012) also reported the prevalence of HBV/HIV co infection to be higher among males (37.5%) than females (24.3%). Vinikoor *et al*. (2015) also demonstrated that HIV co-infected adult males were more likely to be co-infected with HBV than their female counterparts. This finding is also compatible with previous reports from Jos, North Central Nigeria (Irisena et al., 2002), India (Sud et al., 2001) and Zambia (Vinikoor *et al*., 2015). The import of this finding may not be readily discernible (Anigilaje and Olutola, 2013). However, this observation may have been accounted for by the fact that men are more likely to have multiple sex partners and also practice unprotected sex in our polygamous setting (Balogun et al., 2012). The lack of statistically significant difference in this study collaborated with that of Balogun et al. (2012), Anigilaje and Olutola (2013) and Katamba et al. (2020), who reported that HBV presence was independent of gender. On the contrary, Avwioro et al. (2014) reported that females are more infected with HBsAg (5.0%) than males (3.5%) in the same Niger Delta region of Nigeria.

Prevalence of HBV was highest (28.6%) among HIV positive patients aged 6-30 years, and this supported previous studies in Rivers State, Nigeria (Okonko et al., 2020), in North-West Ethiopia (Hou et al., 2005), in Cameroon (Ymele et al., 2012) and in Abuja, Nigeria (Ogundeji, 2018). This may be associated with higher sexual activities within this age group, especially those within adolescent age. Ymele et al. (2012) reported that HBV infection prevalence decreased with the age. Ogundeji (2018) reported that age group of 21-40 years had the predominant HIV, HBV, and HCV prevalence in their study. This finding is in agreement to the finding of Olokoba *et al*. (2011) who reported that women between the ages 25-29 years have a greater prevalence rate. Katamba et al. (2020) also reported that one of the correlates of HIV and hepatitis B co-infections was age (between 20 and 39 years). In all epidemiological studies, younger age has always proved to be the most important factor. The age at which many infections occur calls for concern and concerted efforts aimed at implementing preventive measures that will reduce lifestyle practices among the most susceptible age bracket (Khakhkhar et al., 2012; Ogundeji, 2018). On the contrary, Avwioro et al. (2014) found that individuals within the ages of 31-50 years had the highest prevalence, i.e. HBV infection prevalence increases with the age of the subjects. While Mustapha and Jibrin (2004) reported higher prevalence in age group 41-49 years of age with no case in ≤19 years in Gombe, Nigeria.

In relation to marital status, HBV sero-positivity was higher among HIV-infected individuals who were singles (10.5%) as compared to the married (4.7%) but this was not statistically significant. This may imply that marital status is not really a risk factor for HBV infection but an indicator to consider the sexual partner as a risk for infection, since unmarried people may tend to have many sexual partners or unprotected sex. Also, positivity of HBsAg among the infected married subjects in this study could be an indication that the infection might be through unprotected heterosexual intercourse or close contact with their infected partners as the virus can spread through body fluids. The education-specific prevalence of HIV-HBV co-infections in this study revealed a higher prevalence among those with primary educational status (8.3%) compared to other educational status (tertiary 6.9% and secondary 4.9%). This is compatible with other previous studies. Katamba et al. (2020) also reported that correlates of HIV and hepatitis B co-infections were primary education (especially low level of education), and of course sexual activity. This may be influenced by the settings where our study was conducted, even when majority of the subjects had tertiary education.

Occupational-specific prevalence showed that students had the highest prevalence (22.2%) and closely followed by those in business (16.7%) compared to other occupations, civil servants (7.7%), teaching (6.7%), trading (5.5%) and those with zero prevalence. This might be due to high-risk sexual behaviour among students on campuses. This is not compatible with the low prevalence (12.2%) published for students in a study by Mustapha and Jibrin (2004). This study is also not compatible with the findings of Ikeako et al. (2014) who reported higher prevalence in unemployed subjects and artisans. Ogundeji et al. (2018) also reported higher prevalence of HBV in unemployed subjects and artisans. Ezegbudo et al. (2004) who reported in their study that the occupation of the subjects influenced the infection of antenatal women.

CD4 counts-specific prevalence revealed higher HIV/HBV co-infection rate among HIV-infected individuals with low CD4 cell count <200 cells/μl (15.4%) compared to those with 200-499 cells/μl (6.7%) and ≥500 cells/μl (1.7%). In a related study by Anigilaje and Olutola (2013), many of the children co-infected with HIV-HBV also presented with a much-reduced CD4 counts. In a study by Rawizza et al. (2010), there were no significant differences in the CD4 cell count of children with HIV alone or HIV-HBV-co-infection.

Higher HIV/HBV co-infection was observed among HIV-1 individuals with plasma viral loads 5001 copies/mL and above (13.6%) than those with <40-5000 copies/mL (1.8%). Although this study did not show the effect of HIV-HBV co-infections on HIV replication, it has ben reported previously that HIV-HBV-co-infected subjects have higher HIV RNA levels, are more symptomatic with lower CD4 counts than their counterparts infected with HIV alone (Toussi et al., 2007; Anigilaje and Olutola, 2013).

Although the current study considered only hospital-based cases, it still provides a baseline sero-prevalence data on HIV/HBV co-morbidity. The information obtained from this study will assist policy makers re-direct resources for concomitant screenings/diagnosis of HIV and HBV.

## 4.3 CONCLUSION

The present study has revealed a high prevalence of HBV c-infection in people living with HIV especially among young subjects aged 6–30 years in Uyo, Nigeria. Hence, routine screening of HBsAg should be considered for people living with HIV so as to proffer proper treatment to the co-infected individuals that will improve quality of life and reduce morbidity/mortality.

## Data Availability

ALL DATA ARE CONTAINED IN THE MANUSCRIPT

## ACKNOWLEDGEMENTS

The authors wish to thank the administrations of University of Uyo Teaching Hospital (UUTH), Uyo, Akwa-Ibom State, Nigeria for the ethical approvals and all the individuals who participated in this study. Our immense gratitude also goes to Dr. Emeka Michael, Mr. Inyang Shedrack and Mrs. Onasochi Shedrack at the University of Uyo Teaching Hospital, Uyo, Nigeria.

## REFERENCES

Adoga, M. P., Gyar, S. D., Pechulano, S., Bashayi, O. D., Emiasegen, S. E., Zungwe, T., Iperepolu, O. H., Agupugo, C. and Agwale, S. M. (2010). Hepatitis B virus infection in apparently healthy urban Nigerians: data from pre-vaccination tests. Journal of Infectious Diseases. 4(6):397–400.

Ajayi, B. B., Hamidu, I., Dawurung, J. S., Ballah, A. D. and Isah, J. (2013). Sero-prevalence of some sexually transmitted infectious among antenatal attendees in University of Maiduguri Teaching Hospital. Maiduguri, Nigeria. Annual Biological Research 4(2):141–145

Akwa Ibom State Government official Website gets a new look – The Premium Herald”. The Premium Herald. Retrieved 2016-04-07.

Almeida, P. R. A., Mussi, A. D., Azevedo, S. V. C. and Souto, F. J. (2006). Hepatitis B Virus infection in HIV-positive population in Brazil: results of a survey in the state of Mato Grosso and a comparative analysis with other regions of Brazil. BMC Journal of Infectious Diseases. 6(34): 34

Anggorowati, N., Yano, Y., Heriyanto, D. S., Rinonce, H. T., Utsumi, T. and Mulya, D. P. (2012). Clinical and virological characteristics of hepatitis B or C virus co-infection with HIV in Indonesian patients. Journal of Medical Virology. 84(6): 857–865

Anigilaje EA, Olutola A. Prevalence and Clinical and Immunoviralogical Profile of Human Immunodeficiency Virus-Hepatitis B Co-infection among Children in an Antiretroviral Therapy Programme in Benue State, Nigeria. ISRN Pediatr. 2013 Apr 3;2013:932697. doi: 10.1155/2013/932697.

Aparicio C, Mourez T, Simoneau G, Magnier JD, Galichon B, Plaisance P, et al.(2012). (Proposal of HIV, HBV and HCV targeted screening: short period feasibility study in a freeaccess outpatient medical structure). Presse Med, 41(10): e517–523.

AVERT. 2020. HIV AND AIDS IN NIGERIA. https://www.avert.org/professionals/hiv-around-world/sub-saharan-africa/nigeria. Accessed August 11, 2020

Avwioro OG, Ekene EN, Afadu TE. HIV and HBV co-infection in Niger-Delta, Nigeria. African Journal of Cellular Pathology. 2014;2:48–52.

Baba MM, Gushau W, Hassan AW. Detection of hepatitis-B surface antigenaemia in patients with and without the manifestations of AIDS in Maiduguri, Nigeria. Postgrad Med J. 1998;5(3):125–127.

Babamahmoodi F, Heidari Gorji MA, Mahdi Nasehi M, Delavarian L (2012). The prevalence rate of hepatitis B and hepatitis C co-infection in HIV positive patients in Mazandaran province, Iran. Med Glas Ljek komore Zenickodoboj kantona, 9(2): 299–303.

Balogun TM, Emmanuel S, Ojerinde EF. HIV, Hepatitis B and C viruses’ co-infection among patients in a Nigerian tertiary hospital. Pan Afr Med J. 2012;12:100.

Barth RE, Huijgen Q, Taljaard J, et al.: Hepatitis B/C and HIV in sub-Saharan Africa: an association between highly prevalent infectious diseases. A systematic review and meta-analysis. Int J Infect Dis. 2010; 14(12): e1024–e1031.

Bloquel B, Jeulin H, Burty C, Letranchant L, Rabaud C, Venard V (2014). Occult hepatitis B infection in patients infected with HIV: report of two cases of hepatitis B reactivation and prevalence in a hospital cohort. J Med Virol, 82(2): 206–212.

Bloquel B, Jeulin H, Burty C, Letranchant L, Rabaud C, Venard V (2010). Occult hepatitis B infection in patients infected with HIV: report of two cases of hepatitis B reactivation and prevalence in a hospital cohort. J Med Virol, 82(2): 206–212.

Bloquel, B., Jeulin, H., Burty, C., Letranchant, L., Rabaud, C. and Venard, V. (2010). Occult hepatitis B infection in patients infected with HIV: Report of two cases of hepatitis B reactivation and prevalence in a hospital cohort. Journal of Medical Virology. 82(2): 206–212.

Burnett R. J., G. Francois, M. C. Kew et al., “Hepatitis B virus and human immunodeficiency virus coinfection in sub-Saharan Africa: a call for further investigation,” Liver International, vol. 25, no. 2, pp. 201–213, 2005.

Center for Disease Control and Prevention (CDC, 1997). Revised Guidelines for Performing CD4+ T-Cell Determinations in Persons Infected with Human Immunodeficiency Virus (HIV) MMWR. 1997;46 (no. RR–2):1–29.

Center for Disease Control and Prevention. (2019). Surveillance for acute viral hepatitis. In CDC Surveillance summaries. March 21, 2019 Accessed from; https://www.cdc.int.surveillancesummaries/03.21.00

Chun HM, Roediger MP, Hullsiek KH, et al. Hepatitis B virus co-infection negatively impacts HIV outcomes in HIV sero-converters. J Infect Dis. 2012;205:185–193.

Cooper C. L., E. Mills, B. O. Wabwire, N. Ford, and P. Olupot-Olupot, “Chronic viral hepatitis may diminish the gains of HIV antiretroviral therapy in sub-Saharan Africa,” International Journal of Infectious Diseases, 2009, 13(3): 302–306

Diop-Ndiaye H, Toure-Kane C, Etard JF, Lo G, Diaw P, Ngom-Gueye NF, Gueye NF, et al. Hepatitis B, C sero-prevalence and delta viruses in HIV-1 Senegalese patients at HAART initiation. J Med Virol. 2008 Aug;80(8):1332–6.

Ejele OA, Nwauche C A, Erhabor O. The Prevalence of Hepatitis B surface Antigenaemia in HIV positive patients in the Niger Delta Nigeria. Niger J Med. 2004 Apr-Jun;13(2):175–9.

Ezegbudo CN, Agbonlahor DE, Nwobu GO, Igwe CU, Agba MI, Okpala HO, et al. The sero-prevalence of hepatitis B surface antigen and human immunodeficiency virus among pregnant women in Anambra state, Nigeria. Shiraz E-Med J 2004;5:1–9.

Fernández-Montero Jv, Soriano V. Management of hepatitis C in HIV and/or HBV co-infected patients. Best Pract Res Clin Gastroenterol 2012;26:517–530.

Forbi JC, Gabadi S, Alabi R, Iperepolu HO, Pam CR, Entonu PE, Agwale SM. The role of triple infection with hepatitis Bvirus, hepatitis C virus, and human immunodeficiency virus (HIV) type −1 on CD4+ lymphocyte levels in the highly HIV infected population of North Central Nigeria. Mem Inst Oswaldo Cruz. 2007 Jun;102(4):535–7.

Fujisaki S, Yokomaku Y, Shiino T, Koibuchi T, Hattori J, Ibe S, et al. (2011). Outbreak of infections by hepatitis B virus genotype A and transmission of genetic drug resistance in patients co-infected with HIV-1 in Japan. J Clin Microbiol, 49(3): 1017–1024.

Hoffmann CJ, Seaberg EC, Young S, et al. Hepatitis B and long-term HIV outcomes in co-infected HAART recipients. AIDS. 2009;23:1881–1889.

Hoffmann CJ, Thio CL. Clinical implications of HIV and hepatitis B co-infection in Asia and Africa. Lancet Infect Dis. 2007; 7:402–409

Hou J, Liu Z, Gu F. Epidemiology and prevention of hepatitis B virus infection. Int J Med Sci 2005; 2:50–7.

Ikeako LC, Ezegwui HU, Ajah LO, Dim CC, Okeke TC. Sero-prevalence of human immunodeficiency virus, Hepatitis b, Hepatitis c, syphilis and co-infections among antenatal women in a tertiary institution in south-east Nigeria. Ann Med Health Sci Res 2014; 4:259–63.

Irisena ND, Njoku MD, Idoko JA. Hepatitis surface antigenaemia in patients with human immunodeficiency virus −1(HIV-1) infection in Jos, Nigeria. Nig Med Pract. 2002;41(12):18–20.

Kapembwa KC, Goldman JD, Lakhi S, et al.: HIV, Hepatitis B, and Hepatitis C in Zambia. J Glob Infect Dis. 2011; 3(3): 269–274.

Katamba C, Chungu T and Lusale C. HIV, syphilis and hepatitis B co-infections in Mkushi, Zambia: a cross-sectional study [version 2; peer review: 2 approved]. F1000Research 2020, 8:562 (https://doi.org/10.12688/f1000research.17983.2)

Khakhkhar VM, Bhura PJ, Bhuva SP, Patel CP, Cholera MS. Sero-prevalence of hepatitis B amongst pregnant women attending the antenatal clinic of a tertiary care hospital, Jamnagar (Gujarat). Natl J Med Res 2012; 2:362–5.

Kruse RL, Kramer JR, Tyson GL, Duan Z, Chen L, El-Serag HB, Kanwal F. Clinical outcomes of hepatitis B virus co-infection in a United States cohort of hepatitis C virus-infected patients. Hepatology. 2014 Dec;60(6):1871–8. doi: 10.1002/hep.27337.

Landes M, Newell ML, Barlow P, Fiore S, Malyuta R, Martinelli P, et al. (2008). Hepatitis B or hepatitis C co-infection in HIV-infected pregnant women in Europe. HIV Med, 9(7): 526–534.

Larsen C, Pialoux G, Salmon D, Antona D, Le Strat Y, Piroth L, et al. Prevalence of hepatitis C and hepatitis B infection in the HIV-infected population of France. Euro Surveill. 2008;13(22):18888.

Laurent C, Bourgeois A, Mpoudi-Ngolé E, et al. High rates of active hepatitis B and C co-infections in HIV-1 infected Cameroonian adults initiating antiretroviral therapy. HIV Medicine. 2010;11(1):85– 89.

Lesi O A, Kehinde M O, Oguh DN, Amira CO. Hepatitis B and C virus infection in Nigerian patients with HIV/AIDS. Niger Postgrad Med J. 2007 Jun;14(2):129–33.

Liang HX, Chen YY, Zhou R, Zhang Q, Pan YF, Gu JS, et al. (2010). (A cross-sectional survey of occult hepatitis B virus infection in HIV-infected patients in acquired immune deficiency syndrome area). Zhonghua Shi Yan He Lin Chuang Bing Du Xue Za Zhi, 24(6): 442–444.

Lodenyo H, Schoub B, Ally R, Kairu S, Segal I. Hepatitis B and C virus infections and liver function in AIDS patients at Chris Hani Baragwanath, Johannesburg. East African Medical Journal. 2000;77(1):13–15.

Macfarlane, S.B. (1997). Conducting a Descriptive Survey: 2. Choosing a Sampling Strategy. Tropical Doctors, 27(1):14–21.

Mbanya DN, Takam D, Ndumbet PM. Serological findings amongst first-time blood donors in Yaoundé, Cameroon: is safe donation a reality or a myth? Transfusion Medicine. 2003;13(5):267–273.

Mocroft A, Neuhaus J, Peters L, Ryom L, Bickel M, Grint D, et al. Hepatitis B and C co-infection are independent predictors of progressive kidney disease in HIV-positive, antiretroviral-treated adults. PLoS One 2012;7:e40245.

Moges F, Kebede Y, Kassu A: Sero-prevalence of HIV, Hepatitis B infections and syphilis among street dwellers in Gondar city, Northwest Ethiopia. Ethiop J Health Dev. 2006; 20(3).

Moradi, A., Khodabakhshi, B., Sadeghipour, M., Besharat, S. and Tabarraei, A. (2011). Concurrent infections of hepatitis C and HIV in hepatitis B patients in the north-east of Iran. Tropical doctor. 41(3): 129–131.

Muhammad, H., Adamu, A. S., Ahmad, M. Y., Musa, B., Musa, M. B., Abdulrazaq, G. H. (2013). Prevalence of hepatitis B and C virus infections among HIV-infected patients in a tertiary hospital in North-Western Nigeria. East African Medical Journal. 10(2):76–81.

Mustapha SK and Jibrin YB. (2004). The Prevalence of Hepatitis B Surface Antigenaemia in Patients with Human Immunodeficiency Virus (HIV) Infection in Gombe, Nigeria. Annals of African Medicine, 2004, 3(1): 10–12

Naing, L., Winn, T., and Rusli, B.N. (2006). Practical issues in calculating the sample size for prevalence. Studies Archives of Orofacial Sciences, 1: 9–14.

National Agency for the Control of AIDS (NACA, 2015). GLOBAL AIDS RESPONSE Country Progress Report Nigeria GARPR 2015. National Agency for the Control of AIDS (NACA), Abuja, Federal Republic of NIGERIA. Pp:1–77

National Agency for the Control of AIDS (NACA, 2017). ‘National Strategic Framework on HIV and AIDS: 2017 −2021’. National Agency for the Control of AIDS (NACA), Abuja, Federal Republic of NIGERIA. Accessed August 11, 2020

Nigeria HIV/AIDS Indicator and Impact Survey (NAIIS, 2019). National Agency for the Control of AIDS and the Joint United Nations Programme on AIDS 2019 National HIV Prevalence Report. https://reliefweb.int/report/nigeria/new-survey-results-indicate-nigeria-has-hiv-prevalence-14. Accessed August 10, 2020

Nikolopoulos GK, Paraskevis D, Hatzitheodorou E, Moschidis Z, Sypsa V, Zavitsanos X, et al.(2009). Impact of hepatitis B virus infection on the progression of AIDS and mortality in HIV-infected individuals: a cohort study and meta-analysis. Clin Infect Dis, 48(12): 1763–1771.

Nordenstedt H, White DL, El-Serag HB (2010). The changing pattern of epidemiology in hepatocellular carcinoma. Digestive and Liver Disease, 42: S206–S14.

Nordenstedt, H., White, D. L., & El-Serag, H. B. (2010). The changing pattern of epidemiology in hepatocellular carcinoma. Digestive and liver disease: official journal of the Italian Society of Gastroenterology and the Italian Association for the Study of the Liver, 42 Suppl 3(Suppl 3), S206–S214. https://doi.org/10.1016/S1590-8658(10)60507-5

Nwokedi EE, Epopees MA, Dutse AI. Human immunodeficiency virus and hepatitis B virus co infection among patients in Kano, Nigeria. Niger J Med. 2006 Jul-Sep;15(3):227–9.

Nwolisa E, Mbanefo F, Ezeogu J, Amadi P. Prevalence of hepatitis B co-infection amongst HIV infected children attending a care and treatment centre in Owerri, South-eastern Nigeria. Pan Afr Med J. 2013 Mar 7;14:89.

Ogundeji AA. Sero-prevalence of Human Immunodeficiency Virus, Hepatitis B, Hepatitis C, Syphilis and Co-Infections among Antenatal Women: A Retrospective Case Study at National Hospital Abuja, Federal Capital Territory (FCT), Nigeria. Texila International Journal of Public Health, 2018, 6(2): 1–8

Okonko IO, Adewuyi SA, Omatsone C, Cookey TI. Detection of Hepatitis B Virus Among HIV Positive Fresh Undergraduate Students in Port Harcourt, Nigeria. Asian Journal of Research and Reports in Gastroenterology, 2020, 3(3): 8–13.

Ola SO, Otegbayo GN, Odaibo OD, Olaleye OD, Olubuyide O. Serum Hepatitis C virus and hepatitis B surface antigenaemia in Nigerian patients with acute Icteric hepatitis. West Afr J Med. 2002 Jul-Sep;21(3):215–7.

Olatunji OP, Iseniyi JO. Hepatitis B and C viruses’ co-infection with human immunodeficiency virus infected patients at UITH, Iiorin. Nig Med Pract. 2008;54(1):8–10.

Osborn, M. K., Guest, J. L. and Rimland, D. (2007). Hepatitis B virus and HIV co-infection: relationship of different serological patterns to survival and liver disease. Journal of Medicine. 8(5): 271–279.

Olokoba A B, Salawu F K, Danburam A, Olokoba L B, Midala J K, Badung L H, Olatinwo A. Hepatitis B virus infection amongst pregnant women in North-Eastern Nigeria-A call for action. Niger J Clin Pract 2011;14:10–3.

Opaleye, O., Akanbi, O. and Binuyo, M. (2017). Prevalence of HBV, HIV, and HIV-HBV Co-Infections among Healthcare Workers in Ibadan, Nigeria. BMJ Global Health, 2(Suppl 2): A45

Parboosing R, Paruk I, Lalloo UG. Hepatitis C virus sero-positivity in a South African cohort of HIVinfected, ARV naive patients is associated with renal insufficiency and increased mortality. J Med Virol. 2008 Sep;80(9):1530–6.

Piroth L, Lafon ME, Binquet C, Bertillon P, Gervais A, Lootvoet E, et al. (2008). Occult hepatitis B in HIV-HCV co-infected patients. Scand J Infect Dis, 40(10): 835–839.

Rahimi-Movaghar A, Razaghi EM, Sahimi-Izadian E, Amin-Esmaeili M (2009). HIV, hepatitis C virus, and hepatitis B virus co-infections among injecting drug users in Tehran, Iran. Int J Infect Dis, 14(1): e28–33.

Rahimi-Movaghar, A., Razaghi, E. M., Sahimi-Izadian, E. and Amin-Esmaeili, M. (2009). HIV, hepatitis C virus, and hepatitis B virus co-infections among injecting drug users in Tehran, Iran. International Journal of Infectious Diseases. 14(1): 28–33

Rai, R. R., Mathur, A., Mathur, D., Udawat, H. P., Nepalia, S. and Nijhawan, S. (2007). Prevalence of occult hepatitis B & C in HIV patients infected through sexual transmission. Tropical Gastroenterological Journal. 28(1): 19–23.

Ramezani A, Mohraz M, Aghakhani A, Banifazl M, Eslamifar A, Khadem-Sadegh A, et al. (2009). Frequency of isolated hepatitis B core antibody in HIV-hepatitis C virus co-infected individuals. Int J STD AIDS, 20(5): 336–338.

Rawizza H, Ochigbo S, Chang C, et al. Prevalence of hepatitis co-infection among HIV infected Nigerian children in the Harvard PEPFAR ART program. Proceedings of the 17th Conference on Retroviruses and Opportunistic Infections; February 2010; San Francisco, NC, USA. Poster abstract S–181, http://retroconference.org/2010/Abstracts/39148.htm.

Salmon-Ceron D, Lewden C, Morlat P, et al. Liver disease as a major cause of death among HIV infected patients: role of hepatitis C and B viruses and alcohol. J Hepatol. 2005; 42:799–805.

Schweitzer A, Horn J, Mikolajczyk RT, et al.: Estimations of worldwide prevalence of chronic hepatitis B virus infection: a systematic review of data published between 1965 and 2013. Lancet. 2015; 386(10003): 1546–1555

Shimelis T, Tassachew Y, Tadewos A, et al.: Co-infections with hepatitis B and C virus and syphilis among HIV-infected clients in Southern Ethiopia: a cross-sectional study. HIV AIDS (Auckl). 2017; 9: 203–210.

Stabinski L, O’Connor S, Barnhart M, Kahn RJ, Hamm TE. Prevalence of HIV and Hepatitis B Virus Co-Infection in Sub-Saharan Africa and the Potential Impact and Program Feasibility of Hepatitis B Surface Antigen Screening in Resource-Limited Settings, JAIDS Journal of Acquired Immune Deficiency Syndromes: 2015, 68(p): S274–S285

Stud A, Singh J, Dhiman RK, Wanchu A, Singh S, Chawia Y. Hepatitis B virus co-infection in HIV infected patients. Trop Gastroenterol. 2001;22(2):90–92.

Sungkanuparph S, Vibhagool A, Manosuthi W, Kiertiburanakul S, Atamasirikul K, Aumkhyan A, Thakkinstian A. Prevalence of hepatitis B virus and hepatitis C co infection with human immunodeficiency virus in Thai patients: a tertiary case-based study. J Med Assoc Thai. 2004;87(11):1349–1354.

Teira R; VACH Study Group. Hepatitis-B virus infection predicts mortality of HIV and hepatitis C virus coinfected patients. AIDS 2013; 27:845–848.

Thio CL, Seaberg EC, Skolasky R Jr, et al. HIV-1, hepatitis B virus, and risk of liver-related mortality in the Multicenter Cohort Study (MACS). Lancet. 2002; 360:1921–1926.

Thio CL. Hepatitis B and human immunodeficiency virus co-infection. J Hepatol. 2009;49(5 suppl): S138– S145.

Tounkara, A., Sarro, Y. S., Kristensen, S., Dao, S., Diallo, H. and Diarra, B. (2009). Sero-prevalence of HIV/HBV co-infection in Malian blood donors. Journal of International Association of Physicians AIDS Care. 8(1): 47–51.

Toussi SS, Abadi J, Rosenberg M, Levanon D. Prevalence of hepatitis B and C virus infections in children infected with HIV. Clinical Infectious Diseases. 2007;45(6):795–798

Tsuchiya N, Pathipvanich P, Rojanawiwat A, Wichukchinda N, Koga I, Koga M, et al. (2012). Chronic hepatitis B and C co-infection increased all-cause mortality in HAART-naive HIV patients in northern Thailand. Epidemiol Infect, 1: 1–9.

Vinikoor MJ, Musukuma K, Munamunungu V, et al.: Implementation of routine screening for chronic hepatitis B virus co-infection by HIV clinics in Lusaka, Zambia. HHS Public Access. J Viral Hepat. 2015; 22(10): 858–860.

Wandeler G, Gsponer T, Bihl F, et al. Hepatitis B virus infection is associated with impaired immunological recovery during antiretroviral therapy in the Swiss HIV cohort study. J Infect Dis. 2013; 208:1454–1458.

WHO, Global HIV/AIDS Response: Epidemic Update and Health Sector Progress Towards Universal Access, Progress Report 2011, WHO, Geneva, Switzerland, 2011

World Health Organization (WHO, 2020). HIV and hepatitis co-infections. World Health Organization HIV/AIDS Department, Geneva 27, Switzerland https://www.who.int/hiv/topics/hepatitis/hepatitisinfo/en/

World Health Organization. (2014). HIV/AIDS global and regional statistics report 201 fact sheet. WHO press; Geneva.

World Health Organization. (2015). Guidelines for the prevention, care and treatment of persons with chronic hepatitis B infection. WHO press; Geneva.

World Health Organization: Global health sector response to HIV 2000–2015: focus on innovations in Africa: progress report. Geneva: World Health Organization; 2016.

Ymele FF, Keugoung B, Fouedjio JH, et al. High Rates of Hepatitis B and C and HIV Infections among Blood Donors in Cameroon: A Proposed Blood Screening Algorithm for Blood Donors in Resource-Limited Settings. Journal of Blood Transfusion. 2012; 2012:458372.

